# Prevalence of SARS-CoV-2 antibodies in Denmark 2020: results from nationwide, population-based sero-epidemiological surveys

**DOI:** 10.1101/2021.04.07.21254703

**Authors:** Laura Espenhain, Siri Tribler, Charlotte Sværke Jørgensen, Christian Holm Hansen, Ute Wolff Sönksen, Steen Ethelberg

## Abstract

**Background:** Seroprevalence studies have proven an important tool to monitor the progression of the coronavirus disease 2019 (COVID-19) epidemic. We present results of consecutive population-based seroprevalence surveys performed in Denmark in 2020.

**Methods:** Invitation letters including a questionnaire covering symptoms were sent to representatively drawn samples of the population in spring, late summer and autumn/winter of 2020. Blood samples from participants taken at public test-centers were analyzed for total Ig and seroprevalence estimates per population segment calculated and compared to other surveillance parameters.

**Results:** From 34,081 participating individuals (response rate 33%), we obtained seroprevalence estimates increasing from 1.1% (95%CI: 0.7%–1.7) in May to 4.0 % (95%CI: 3.4%–4.7%) in December 2020. By December 2020, 1.5% of the population 12 years and older had tested positive by PCR. Seroprevalence estimates were roughly 3 times higher in those aged 12-29 compared to 65+ and higher in metropolitan municipalities. Among seropositives, loss of taste/smell were the more specific symptoms, 32%-56% did not report any symptoms. In half of seroconverted families, we did not see evidence of transmission between generations. Infected individuals in older age groups were hospitalized several fold more often than in younger.

**Conclusions:** Seroprevalence increased during 2020; younger age groups were primarily infected in the autumn/winter surge. Approximately half were asymptomatically infected. Denmark has a high per capita test rate; roughly two undiagnosed infections of COVID-19 were estimated to occur for each diagnosed case. The epidemic appears to have progressed relatively modestly during 2020 in Denmark.

**summary:** We describe population-based COVID-19 seroprevalence surveys performed in Denmark in 2020. The seroprevalence increased during the year, particularly in adolescents and young adults, but was overall low. Roughly two undiagnosed cases per PCR-confirmed case were detected by December 2020.

## BACKGROUND

The degree to which the COVID-19 pandemic is spreading through different countries or regions may be assessed through population-based seroprevalence studies, which aim to quantify the proportion of the population that has developed antibodies against SARS-CoV-2. Such studies have to date been performed in a number of countries [1-9].

Similar to several other European countries, Denmark experienced increased transmission of SARS-CoV-2 infection in spring and late autumn 2020. A comprehensive lock-down was imposed in March 2020, gradually lifted towards summer and again gradually reintroduced during autumn and winter [10]. The Danish National Seroprevalence Survey of SARS-CoV-2 infection (DSS) was initiated in the spring of 2020, following a parliamentary decision in April 2020, which called for a representative population study to be performed. The study design was set by recommendation from a group of independently appointed national experts in April 2020 [11] and seroprevalence subsequently determined at several time points throughout 2020. Here we describe the set-up and results from DSS and relate the results to the national surveillance of PCR diagnosed COVID-19-cases.

## METHODS

### Design and study population

DSS is a nationwide population-based prevalence survey aiming to investigate seroprevalence for SARS-CoV-2. The study was launched in the spring of 2020 and performed by Statens Serum Institut (SSI) over three rounds: In May 2020 (DSS-I), August 2020 (DSS-II) and September-December 2020 (DSS-III).

For each survey round, a random population sample of Danish residents was drawn from the Danish civil registry [12]. For DSS-I, adults aged 18 years and older living in one of 30 municipalities which had a test facility (see below) at that time (n=5), or were neighboring a municipality with a test facility (n=25) were eligible (approximately 45% of the population of Denmark). For DSS-II and DSS-III, people aged 12 years or older were selected by random sampling, with no restriction on municipality (n=98). Parents living on the same address as invited children 12-17 years old, were also invited to participate.

### Recruitment

We primarily invited participants using the Danish digitalized postal system covering 90% of the Danish population [13]. Invitation letters (as pdf’s) were sent via the secure, digital mailbox-system (“e-Boks”), using the civil registration number. Additionally, physical letters were sent by regular mail to invitees below 18 years of age and to those without e-Boks. For DSS-I we invited 2,600 people on May 5 and 15. For DSS-II we invited 6,000 people on August 15, 21 and 28. For DSS-III we invited 70,000 people over a 14 week period from September 11 to December 11, 2020. Letters of invitation contained information about: the aim and study design, details about the antibody test (how to interpret and understand the test result), and how to book a test. DSS-II and DSS-III also included a link to an electronic questionnaire. Invitations and questionnaires were available in Danish, English and Arabic language versions. The questionnaire contained, among others, questions about current and past symptoms.

### TestCenter Denmark, sample collection and analysis

Blood sampling was performed at test stations of ‘TestCenter Denmark’, a public national SARS-CoV-2 test facility system established during March and April 2020 [14, 15]. At nation-wide facilities it offers free of charge easy access PCR testing for asymptomatic or mildly symptomatic individuals, in addition to the existing laboratories at the acute care hospitals that test symptomatic individuals and hospitalized patients upon admission. Timeslots for PCR testing were booked by the individual through a secure website. By May 2020, 22 PCR-test facilities had been established, of which five facilities were additionally equipped for taking blood samples. DSS-I made use of these five test stations. By August 2020, antibody testing could be carried out in 17 of the 22 PCR-test facilities. These were used for DSS-II and DSS-III. Study participants could book timeslots for antibody testing using the same IT-platform as for PCR testing. Transportation to a test facility was at participants’ own cost. Blood sampling was performed by medically trained personnel. Five ml blood samples were taken in BD Vacutainer® Serum tubes. The sample was packed, collected and transported to SSI for analysis. Total serum concentration of anti-SARS-CoV-2 immunoglobulin was measured by use of the Wantai SARS-CoV-2 Ab ELISA (Beijing Wantai Biological Pharmacy Enterprise, Beijing, China) according to the manufacturer’s instructions.

### Data source and the Danish COVID-19 surveillance data

The Danish Microbiological Database (MiBa) contains all microbiological test results from all clinical microbiological departments in Denmark and microbiological and serological results from TestCenter Denmark [16]. Using MiBa we identified antibody test results and previous COVID-19-PCR-test results amongst the study participants. We used information on number of admitted and deceased by date, sex and age group from the Danish surveillance system of COVID-19 [17, 18]. This involves daily registry linkage to the National Patient Registry [19] for information about hospital admissions and to the Civil Registry and The National Cause of Death Register [20] for information about deaths within 30 days for PCR-diagnosed COVID-19 cases.

### Statistical analyses

We included all persons with a conclusive antibody test result within ten weeks from the invitation. We estimated the seroprevalence as the proportion of participating individuals with a positive antibody test result. We adjusted the seroprevalence estimates for test sensitivity (0.97) and specificity (0.995) (internal assessment at SSI) using the Rogan-Gladen estimator [21] and computed 95% confidence intervals using Blaker’s method [22]. We present seroprevalence by age group, sex, the five geographical regions of Denmark and classification of municipality (capital, metropolitan, provincial, commuter, rural) as defined by Statistics Denmark [23]. To evaluate whether the variation in response rate by age groups, sex and region affected the estimated seroprevalence, we predicted the value (0 = negative and 1 = positive) of the missing serology test by multiple imputation including sex, age group and region in the prediction model.

For the analyses, we defined four periods (May, August, October and December) as the time periods the seroprevalence estimate reflected. We compared the estimates of infected individuals to the number of PCR test positive, hospitalized and deceased in national surveillance. To do that, we subtracted 14 days from the mean date of blood sampling in the four periods to compare with PCR-test positive from the national surveillance system, and added 10 days to find the comparable date for hospital admission and 20 days for number of deaths.

### Ethical and legal considerations

The DSS was performed as a national disease surveillance project, registered with the Danish Data Protection Agency and approved regarding legal, ethical and cyber-security issues by the SSI Compliance department in conjunction with the Danish governmental law firm. Participation in the survey was voluntary and invitees received information about the selection procedure, risks associated with participation, data security issues, their legal rights, including the right to withdraw from the study, and the use of their data and results in the letter of invitation.

## RESULTS

### Participation

The COVID-19 incidence and test intensity in Denmark in 2020 is depicted in Figure 1. The three DSS study rounds had 2,512 (48%), 7,015 (39%), and 18.161 (26%) participants, respectively (Table 1). The median dates of sampling for the four defined study periods were May 18 (defined as ‘May’), September 19 (‘August’), November 6 (‘October’), and December 16 (‘December’) (Figure 1).

**Table 1.** Number of invited persons and proportion who participated by DSS, age group, sex, and region.

| Group | DSS-I* |  | DSS-II |  | DSS-III |  |
| --- | --- | --- | --- | --- | --- | --- |
|  | Invited | Participation | Invited | Participated | Invited | Participated |
| Total | 5,200 | (48) | 18,000 | (39) | 70,000 | (26) |
| Age group |  |  |  |  |  |  |
| 12-17 years |  |  | 1,492 | (31) | 5,631 | (20) |
| 18-39 years | 2,146 | (40) | 5,715 | (32) | 22,105 | (22) |
| 40-64 years | 2,077 | (56) | 6,700 | (48) | 26,173 | (33) |
| 65+ years | 977 | (50) | 4,093 | (36) | 16,091 | (22) |
| Sex |  |  |  |  |  |  |
| Female | 2,585 | (53) | 9,132 | (44) | 35,282 | (29) |
| Male | 2,615 | (44) | 8,868 | (34) | 34,718 | (23) |
| Region |  |  |  |  |  |  |
| Capital | 2,167 | (48) | 5,680 | (42) | 22,268 | (30) |
| Zealand | 619 | (43) | 2,618 | (39) | 10,107 | (26) |
| Southern Denmark | 798 | (46) | 3,737 | (35) | 14,646 | (20) |
| Mid Jutland | 1,035 | (52) | 4,108 | (38) | 15,865 | (23) |
| North Jutland | 581 | (52) | 1,857 | (40) | 7,113 | (29) |

\*Includes only people 18 years and older living in one of 30 municipalities which had a test facility at the time (n=5) or
was neighboring a municipality with a test facility (n=25). Approximately 45% of the population of Denmark

**Figure 1.**
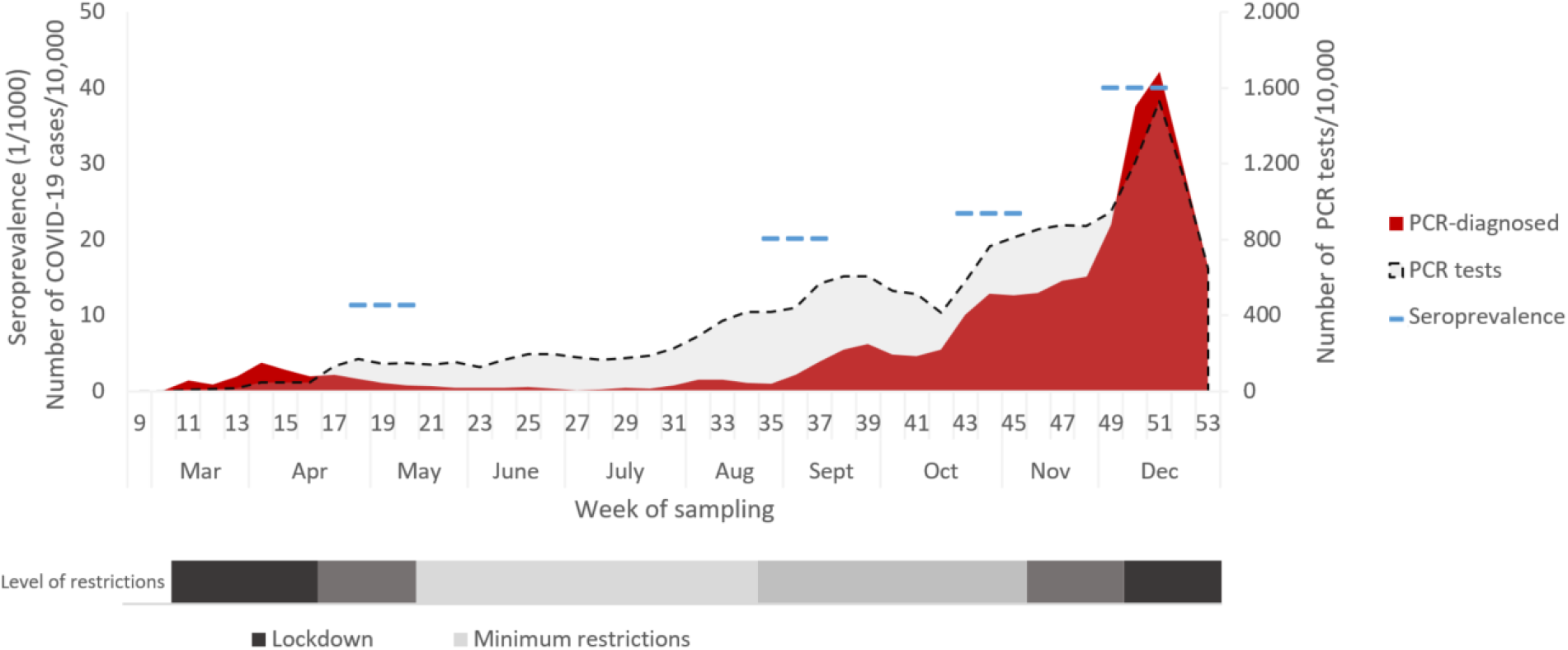
Number of cases (per 10,000), PCR tests (per 10,000), seroprevalence (per 1,000) and measures to reduce transmission by week, Denmark 2020

Overall participation was lower in males and younger age groups (Table 1). For DSS-II and DSS-III respectively, the questionnaire was filled in by 2,737 (39%) and 10,358 (57%) of the participants.

### Seroprevalence

The proportion of participants with detectable SARS-CoV-2 antibodies increased from 1.1% (95%CI: 0.7%–1.7%) in May 2020 to 4.0 % (95%CI: 3.4%–4.7%) in December 2020 (Table 2). Restricting the analysis to match the narrower geographic and age inclusion criteria for DSS-I, the estimated seroprevalence in December was 5.1% (95%CI 4.0%–6.2%). When taking the non-response by age group, sex and region of residence into account by imputation, the estimates increased with up to 0.4 percentage points (Table 2). Point estimates tended to be higher in the two younger age groups (12 to 17 years and 18 to 39 years of age), lower in the 65 years and older age group (Figure 2), and higher in the Capital region than in the other four regions. No difference was observed by sex (Table 2).

**Table 2.** Seroprevalence of SARS-CoV-2 in May, August – December 2020, by age group, sex, region and type of municipality.

|  | May* (n=2,512) |  | August (n=11,478) |  | October (n=9,654) |  | December (n=4,044) |  |
| --- | --- | --- | --- | --- | --- | --- | --- | --- |
|  | % | (95% CI) | % | (95% CI) | % | (95% CI) | % | (95% CI) |
| Total | 1.1 | (0.7 - 1.7) | 2.0 | (1.7 - 2.3) | 2.2 | (1.9 - 2.6) | 4.0 | (3.4 - 4.7) |
| Adjusted total | 1.5 | (0.9 - 2.1) | 2.0 | (1.7 - 2.4) | 2.4 | (1.9 - 2.8) | 4.3 | (3.4 - 5.1) |
| Age group |  |  |  |  |  |  |  |  |
| 12-17 years |  |  | 0.9 | (0.2 - 2.2) | 2.8 | (1.6 - 4.5) | 6.4 | (3.8 - 10) |
| 18-39 years | 2.3 | (1.1 - 3.6) | 2.8 | (2.2 - 3.6) | 3.3 | (2.6 - 4.1) | 5.2 | (4.0 - 6.6) |
| 40-64 years | 0.6 | (0.0 - 1.1) | 2.0 | (1.5 - 2.4) | 2.0 | (1.5 - 2.5) | 3.5 | (2.7 - 4.6) |
| 65+ years | 0.5 | (0.0 - 1.1) | 1.5 | (0.9 - 2.1) | 1.1 | (0.6 - 1.8) | 2.3 | (1.2 - 3.8) |
| Sex |  |  |  |  |  |  |  |  |
| Female | 0.8 | (0.3 - 1.6) | 2.1 | (1.7 - 2.5) | 2.0 | (1.6 - 2.4) | 4.1 | (3.3 - 5.1) |
| Male | 1.5 | (0.7 - 2.5) | 2.0 | (1.5 - 2.4) | 2.6 | (2.0 - 3.1) | 3.9 | (3.0 - 5.1) |
| Region |  |  |  |  |  |  |  |  |
| Capital | 1.8 | (1.0 - 2.9) | 3.2 | (2.6 - 3.8) | 3.2 | (2.6 - 3.9) | 5.0 | (3.9 - 6.2) |
| Zealand | 1.8 | (0.5 - 4.4) | 1.8 | (1.1 - 2.7) | 1.3 | (0.7 - 2.1) | 3.9 | (2.4 - 6.0) |
| South | 0.6 | (0 - 2.3) | 1.5 | (0.9 - 2.2) | 1.6 | (0.9 - 2.4) | 3.2 | (2.0 - 4.9) |
| Mid Jutland | 0.2 | (0 - 1.4) | 1.2 | (0.7 - 1.8) | 2.0 | (1.3 - 2.7) | 3.5 | (2.1 - 5.1) |
| North Jutland | 0.5 | (0 - 2.4) | 1.1 | (0.5 - 2.0) | 1.6 | (0.8 - 2.5) | 3.0 | (1.4 - 5.3) |
| Type of area (municipality) |  |  |  |  |  |  |  |  |
| Capital | 1.8 | (1.0 - 2.9) | 3.2 | (2.6 - 4.0) | 3.5 | (2.8 - 4.2) | 5.6 | (4.4 - 7.0) |
| Metropolitan | 0.8 | (0.2 - 1.9) | 1.8 | (1.1 - 2.6) | 1.5 | (0.7 - 2.3) | 4.9 | (3.2 - 7.0) |
| Provincial | 3.1 | (0.9 - 7.6) | 1.5 | (0.9 - 2.1) | 2.2 | (1.6 - 3.0) | 3.5 | (2.3 - 5.0) |
| Commuter | 0 | (0 - 1.1) | 1.9 | (1.2 - 2.7) | 0.7 | (0.2 - 1.5) | 3.3 | (1.9 - 5.1) |
| Rural | 0 | (0 - 2.7) | 0.9 | (0.5 - 1.6) | 1.7 | (1.0 - 2.5) | 1.2 | (0.4 - 2.7) |

\*Includes only people 18 years and older living in one of 30 municipalities which had a test facility at the time (n=5) or was neighboring a municipality with a test facility (n=25). Approximately 45% of the population of Denmark.

**Figure 2.**
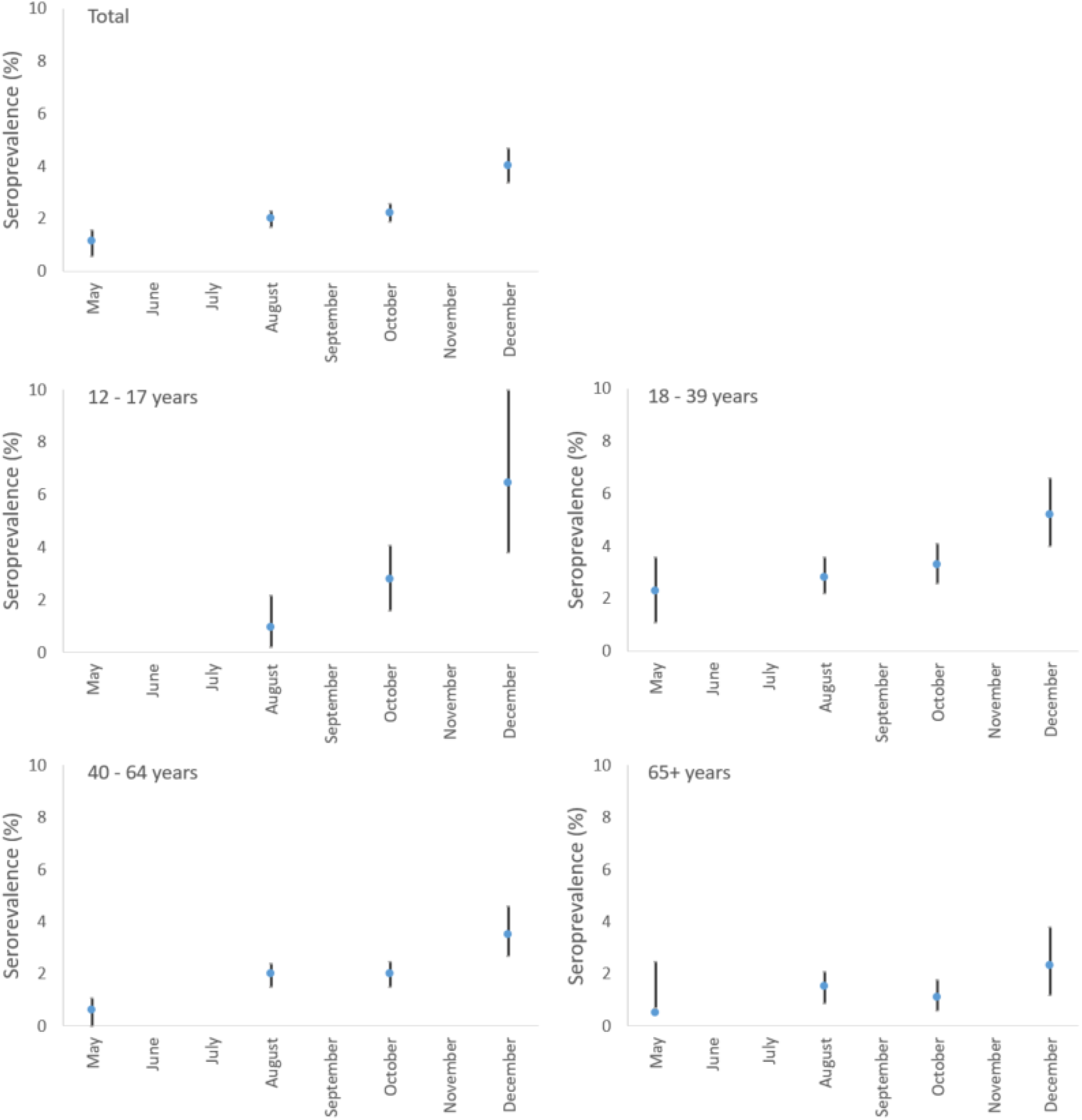
Seroprevalence of SARS-CoV-2 in May, August — December 2020, by age group, Denmark

### Seroprevalence of children and their parents

A total of 1,244 families had a child and at least one parent tested. Among these, 79 (6.4%) families were found to have at least one seropositive family member. These included 3.2% with a seropositive child and 4.2% families with at least one seropositive parent. In 21 of the 79 seropositive families (27%), both child and parent(s) were seropositive, in 19 families (24%) only the child was seropositive and in 39 (49%) seropositive families the child was seronegative.

### Symptoms among seropositive participants

Of the 369 seropositive participants who filled in the questionnaire, 59% reported having experienced at least one of five core symptoms (fever, cough, shortness of breath and/or loss of sense of taste or smell) since February 2020, versus 28% among seronegative participants (Table 3). Loss of smell or taste, reported by 24% and 23% of the seropositives, respectively, were highly associated with seropositivity (odds ratios of 16 and 14, respectively, Table 3).

**Table 3.** Number, proportion (%) of participants and odds ratio of symptoms, by antibody status, DSS-II and DSS-III, Denmark 2020.

| Symptom* | Antibody test result |  |  |  |  |  |
| --- | --- | --- | --- | --- | --- | --- |
|  | Positive (n=369) |  | Negative (n=12,726) |  | Odds ratio | 95%CI |
|  | Number | Proportion (%) | Number | Proportion (%) |  |  |
| No symptoms reported | 112 | (30) | 7,232 | (57) | 0.33 | (0.27-0.42) |
| At least one relevant symptom** | 216 | (59) | 3,508 | (28) | 3.7 | (3.0-4.6) |
| Fever | 139 | (38) | 1,801 | (14) | 3.7 | (3.0-4.6) |
| Cough | 141 | (38) | 2,544 | (20) | 2.5 | (2.0-3.1) |
| Shortness of breath | 71 | (20) | 811 | (6) | 3.6 | (2.7-4.7) |
| Loss of sense of smell | 88 | (23) | 241 | (2) | 16 | (12-21) |
| Loss of sense of taste | 84 | (22) | 263 | (2) | 14 | (11-19) |
| Other symptoms*** | 41 | (11) | 1,986 | (16) | 0.68 | (0.47-0.94) |

\* In the period since 1 February 2020
\*\*Relevant symptoms includes one or more of the following: fever, cough, shortness of breath and/or loss of sense of taste or smell
\*\*\* Muscle ache, eye pain, head ache, colored sputum, runny nose, sneezing, back pain, tiredness without one of the five relevant symptoms

In DSS-II 44% (95%CI: 32%-56%) of the seropositive participants reported no symptoms since February and a further 9% reported symptoms not belonging to the five mentioned core symptoms. In DSS-III the equivalent figures were 27% and 11%.

### Previous PCR positive participants

Of the altogether 104,413 invited persons (93,200 randomly selected Danish residents and 11,213 parents to selected children), 871 had a positive PCR-test result prior to the antibody test or prior to the invitation to the study. Of these, 271 (31%) had an antibody test taken 12 days or more after the positive PCR result. SARS-CoV-2 antibodies were detected in 251 of the previous PCR positive persons (95% when adjusting for the sensitivity and specificity of the test) and 20 did not have detectable antibodies. The median time in days between the positive PCR result and the positive antibody test result was 56 days range [12–293] and 59.5 days range [13–213] for the 20 seronegative persons.

### Seroprevalence in relation to PCR-diagnosed, admitted and deceased COVID-19 cases

According to the national COVID-19 surveillance system, 78,125 persons above 12 years (153/10,000 inhabitants) had tested positive for SARS-CoV-2 by PCR in Denmark by December 2 (Figure 1). Our finding of a seroprevalence of 4.0% (95%CI 3.4–4.7) corresponds to a total of 205,000 (172,000–239,000) persons above 12 years of age having been infected in Denmark as by December 2020. Thus the estimated ratio of infected to PCR-diagnosed cases was three in December and six in May 2020 (Table 4). The estimated ratio varied by age. It was higher in the 18-39 year age group in May and August and decreased during autumn. No obvious pattern was seen for the 65-year and older age group, by region or sex during the period (Table 4). The infection fatality rate and rate of admitted out of the estimated number of infected increased markedly with older age (Table 4).

**Table 4.** Ratio between estimated number of infected and PCR-diagnosed cases from national surveillance at four time points, number of COVID-19 admissions, infection admission rate, number of COVID-19 deaths and infection fatality rate by December 2020, by age group and sex, Denmark 2020.

|  | Estimated ratio of infected/PCR-diagnosed COVID-19 cases |  |  |  | Estimated number of infected by December 2 | Admissions per December 12** | IAR (%) | Death per December 22** | IFR (%) |
| --- | --- | --- | --- | --- | --- | --- | --- | --- | --- |
|  | May* | August | October | December |  |  |  |  |  |
| Total | 6 | 6 | 3 | 3 | 205,000 | 5,987 | 3.0 | 1,081 | 0.53 |
| Age group |  |  |  |  |  |  |  |  |  |
| 12-17 years | - | 5 | 4 | 3 | 26,000 | 58 | 0.22 | 0 | - |
| 18-39 years | 13 | 7 | 4 | 2 | 83,000 | 642 | 0.77 | 3 | 0.00 |
| 40-64 years | 2 | 5 | 3 | 2 | 67,000 | 2,033 | 3.0 | 56 | 0.08 |
| 65+ years | 3 | 5 | 3 | 3 | 27,000 | 3,164 | 12 | 1,022 | 3.8 |
| Sex |  |  |  |  |  |  |  |  |  |
| Female | 4 | 6 | 3 | 3 | 105,000 | 2,790 | 2.6 | 480 | 0.46 |
| Male | 9 | 6 | 4 | 3 | 100,000 | 3,107 | 3.1 | 601 | 0.60 |

\*Includes only people 18 years and older living in one of 30 municipalities which had a test facility at the time (n=5) or was neighboring a municipality with a test facility (n=25). Approximately 45% of the population of Denmark
\*\*Among confirmed COVID-19 cases from national surveillance
IAR=Infection admission rate. IFR=Infection fatality rate

## DISCUSSION

In this national representative seroprevalence study among Danish residents aged 12 years and older, we found detectable SARS-CoV-2 antibodies in 4.0 % (95%CI: 3.4–4.7%) of the participants by the beginning of December 2020. This was four times more than the estimate from May and twice the estimate of August 2020. This study also provides information on the regional and demographic progression of the epidemic and its results can be interpreted in the context of other surveillance parameters.

The seroprevalence varied by geography and age group, which is consistent with the general picture from the national surveillance system. It appears that people from more densely populated urban areas were infected in the early stage of the epidemic, and that the epidemic only gradually spread to the less densely populated areas later. Our results are in line with the serological surveys of blood donors which has also been carried out in Denmark [24], although this group may not be representative of the general population.

Between May and December 2020, PCR testing rose from 150 to more than 1000 tests per 10,000 inhabitants per week. Our results show that the proportion of PCR-confirmed COVID-19 cases out of all estimated infections was 1:5 in the spring but just 1:2 by early December. There were relatively more undiagnosed cases in the 18-39-year age group during the first six month of the epidemic, suggesting that this age group may experience a less severe course of disease. It is well established that severity of illness increases markedly with age [25] and the estimated infection fatality and infection admission rate also increased markedly by age group in this study.

National representative seroprevalence surveys from other countries have shown quite variable seroprevalence estimates [26]: Surveys in France [27] in May, and Spain [2] and Brazil [9] in June and the Netherlands [7] in July 2020 estimated that respectively around 4.5%, 5%, 3.5% and 4% of the population had been infected with SARS-CoV-2 at that point. All four surveys revealed substantial geographical variability. In September a survey carried out in the US found that in 25 of 49 jurisdictions with sufficient samples to estimate seroprevalence more than 5% of people had detectable SARS-CoV-2 antibodies [1]. Our results are comparatively low and thus the epidemic appears to have affected Denmark only mildly in 2020. This may be corroborated by the cumulated mortality numbers, which by 31 Dec 2020 were 23 per 100,000 [17], placing Denmark in the lower end of the European scale [28].

Estimates from other studies of the share of asymptomatic infections out of the total number of SARS-CoV-2 infections vary notably from a few percent to 41% with a pooled overall proportion of 17% found in a recent meta-analysis [29-31]. In DSS-II carried out in the late summer, 44% of the seropositive participants did not recall having had any symptoms of acute infection since February 2020 and an additional 9% reported symptoms not typically associated with COVID-19. The percentage reporting no symptoms since February 2020 fell to 27% in the DSS-III. However at this point other respiratory illnesses may increasingly have affected the results. Thus, our best estimate is that around half of the seropositive persons had an asymptomatic infection. We found that loss of smell or taste, experienced by almost ¼ with SARS-CoV-2 antibodies, were by far the more specific symptoms of COVID-19 infection.

In more than two thirds of families with at least one seropositive family member, only the parent(s) or the child had seroconverted, indicating that transmission between generations within households is the exception rather than the rule. We were unable to disentangle the chain of transmission between generations.

Though actual numbers were low, we found that <5% of previous PCR positive participants did not have detectable SARS-CoV-2-antibodies 12 days after their first positive PCR test. This might be because of waning immunity, or that the individuals did not elicit a detectable antibody response (possibly due to mild or asymptomatic infection). The proportion does correspond to what has been reported from Iceland [32] and a study in preprint from the UK [6].

Denmark has a relatively high degree of IT penetration and frequently makes use of national registers and public digital resources. Utilization hereof was among the strengths of this study. From the Danish national civil register, it was possible to obtain a random sample of residents, and identify the parents of those below 18 years of age. Individually referable national surveillance data allowed us to identify all previous PCR tests amongst the participants and relate this to their antibody-status. Another strength of the study was the use of the logistical set-up of the large state-driven test-system, TestCenter Denmark, that was created as a parallel system to the existing clinical test system located at the hospitals. Using the existing set up for PCR testing, meant that most people had an easy access to a test facility for antibody testing. However, due to the geographical distribution of the test stations, some persons had quite long driving distances to a test facility (up to 100 km), which may have affected their willingness to participate in the study.

The study had several limitations. When interpreting the findings, the suboptimal participation rates should be taken into account. Participation decreased from 48% in DSS-I, through 39% in DSS-II to 26% in DSS-III, and even fewer replied to the questionnaire concerning symptoms. Even though the drawn sample is representative of the population, participation may not be. We do not know if certain subgroups of the population were underrepresented in the study, but may presume that hard to reach populations would be so. The seroprevalence estimates were stable but slightly underestimated when taking the non-response by age group, sex and region of residence into account.

In conclusion, our study provides estimates of SARS-CoV-2 dissemination in Denmark at four time points, based on a representative sample of the population and relate it to the number of PCR-confirmed COVID-19 cases in the national surveillance system. We found that the epidemic had predominantly affected the capital and metropolitan areas and saw indications of a higher seroprevalence in young adults throughout the epidemic, although children 12-17 years old were mainly affected in the second surge of the epidemic. Overall, the estimated seroprevalence in Denmark throughout 2020 was low, compared to other countries. The results seem to support that the measures introduced in Denmark in the spring of 2020 and onwards have been effective in keeping the epidemic from developing rapidly in the community, however also indicate that the majority of the population is still at risk of contracting COVID-19. As more than one in three infections seem to be asymptomatic, social distance measures and efforts to identify and isolate new cases and their contacts are imperative for future epidemic control.

## Data Availability

Data is not publicly available

## Notes

### Author contributions

Study conception and design: LE, ST and SE. Acquisition of data: all authors. Analysis and interpretation of data: all authors. LE, ST and SE drafted the first version of the article, all authors took part in revising it critically for important intellectual content and approved of the final version to be published. LE and CHH had full access to all of the data and take responsibility for the accuracy of the data analysis.

### Disclaimer

The funder asked for a ‘population representative study design’. The funder had no role in data collection and analysis, writing of or decision to submit the manuscript for publication.

## Acknowledgements

We thank everyone who participated in this study by giving blood. We thank all staff in TestCenter Denmark, including staff in test stations, the involved SSI departments and the expert group that advised on the design of the study.

